# Examination of Influenza A Infection Rate, Its Determinants, and Seasonal Influenza Vaccine Effectiveness in the Post-COVID-19 Pandemic Era

**DOI:** 10.1101/2023.07.08.23292128

**Authors:** Isabell Wagenhäuser, Juliane Mees, Julia Reusch, Thiên-Trí Lâm, Alexandra Schubert-Unkmeir, Lukas B. Krone, Anna Frey, Oliver Kurzai, Stefan Frantz, Lars Dölken, Johannes Liese, Alexander Gabel, Nils Petri, Manuel Krone

**Author notes:** Corresponding author: Isabell Wagenhäuser, Infection Control and Antimicrobial Stewardship Unit, University Hospital Würzburg, Josef-Schneider-Str. 2 / E1, 97080 Würzburg, Germany. both authors contributed equally (Nils Petri, Manuel Krone).

## Abstract

In the context of the COVID-19 pandemic, a pronounced wave of Influenza A occurred in the 2022/23 winter season under generally relaxed post-pandemic non-pharmaceutical preventive measures.

**Aim:** This study aimed to investigate the Influenza A infection rate, factors influencing its occurrence and seasonal Influenza vaccine effectiveness on seroconversion in the post-COVID-19 pandemic era.

**Methods:** The seroconversion of Anti-Influenza-A-Nucleoprotein/Matrix IgG was investigated in 402 healthcare workers (HCWs) during the winter season of 2022/2023 (23 May 2022 to 11 May 2023). The participants provided a serum sample and completed a study questionnaire both before and after the seasonal Influenza A wave (24 October 2022 to 8 January 2023). The levels of a vaccine-independent Anti-Influenza-A-Nucleoprotein/Matrix IgG were measured using the SERION ELISA *classic* Influenza A IgG assay, with a 2-fold increase as indicative of seroconversion after asymptomatic or symptomatic influenza infection.

**Results:** Among the participants, 20.6% (95% CI 17.0-24.9%; 83/402) showed seroconversion. The multivariate logistic regression analysis revealed that the age category of ≥ 45 years (p=0.03) and regular patient contact (p=0.02) significantly influenced seroconversion. However, the factors male gender, BMI, smoking, household size, seasonal Influenza vaccination, and SARS-CoV-2 infection during the Influenza A season were not significantly associated with seroconversion. The effectiveness of the 2022/23 seasonal Influenza vaccine on seroconversion induced by Influenza infection was 22.6% (95% CI -17.1-50.6%).

**Conclusion:** During the initial Influenza A season following the COVID-19 pandemic, approximately 20% of HCWs contracted an Influenza A infection. This highlights a potential risk and a significant asymptomatic or symptomatic infection rate posing a theoretical risk for intrahospital transmission chains and nosocomial infections.

## 1 Introduction

The Influenza virus is a significant respiratory pathogen that poses a considerable burden in terms of morbidity and mortality. (1, 2) Following the COVID-19 pandemic, which witnessed minimal Influenza infection waves in the 2020/21 and 2021/22 winter seasons, there were two distinct Influenza waves during the 2022/23 winter season in Germany. In late autumn, there was an Influenza A wave affecting the general population, and in spring 2023, Influenza B predominantly affected children and adolescents. These waves occurred under reduced measures within the framework of the expiring COVID-19 pandemic. (3, 4)

The importance of Influenza A and its seasonal increase applies especially to healthcare workers (HCWs) who are at higher risk of contracting Influenza from infected patients. (5) It is crucial to consider the risk of nosocomial infections and the role of HCWs as key players in intrahospital transmission chains, especially considering that Influenza A is a central viral pathogen causing acute respiratory infections (ARI) and possesses significant morbidity and pandemic potential compared to other human pathogenic Influenza viruses. (6, 7) Consequently, it is essential to establish and enhance optimal Influenza A prevention strategies for HCWs, with seasonal vaccination being one recommended approach. (8, 9) In view of the critical infrastructure of hospitals, it is essential in this context to minimise the inability to work due to Influenza A infection through prevention. (10) The important prerequisite for the development of prevention strategies is the best possible evaluation of the initial situation and thus of the epidemiology and the Influenza A infection rate defined as the share of a population contracting Influenza A during a season. (11, 12)

Therefore, it is crucial to assess the Influenza A infection rate among HCWs during the Influenza season. This evaluation should account for seroconversion, which includes unreported infections that were asymptomatic or not virologically diagnosed. Additionally, it is important to identify the various factors influencing Influenza infection, primarily the effectiveness of seasonal Influenza vaccination. (2, 13, 14)

However, current evidence in this field is limited due to the evaluation of immunocompromised or chronically ill individuals, (15) and, most importantly, due to analyses in study periods prior to the COVID-19 pandemic. (16, 17, 18) The COVID-19 pandemic has significantly changed seasonal vaccination preparedness in recent years, (19) as well as various hygiene measures, (20) most notably the establishment of face masks, (21) which limits the applicability of previous studies to the present. Consequently, previous studies may not fully apply to the present context, thus leaving a gap in understanding the Influenza A infection rate and its influencing factors among HCWs in the post-COVID-19 pandemic era.

This study aims to evaluate the Influenza A infection rate among HCWs during the 2022/23 Influenza A season and its determinants, including the vaccine effectiveness of the seasonal tetravalent Influenza vaccine for the 2022/23 season.

## 2 Methods

### 2.1 Study setting

This study was conducted as a substudy of the CoVacSer cohort study which prospectively investigates the SARS-CoV-2 immunity, quality of life, and ability to work in HCWs after COVID-19 vaccination and/or SARS-CoV-2 infection. Inclusion criteria for study enrolment were: (i) age ≥ 18 years, (ii) written consent form, (iii) employment in healthcare sector, and (iv) history of SARS-CoV-2 infection and/or COVID-19 vaccination with an EMA approved vaccine. The data presented was collected and included independently of the subjects’ COVID-19 vaccination and SARS-CoV-2 infection history. In the case of a seasonal Influenza vaccination, the study protocol included an additional participation from 14 days after the Influenza vaccination. As seasonal Influenza vaccination, which is recommended as occupational indication for HCWs in Germany, (9) predominantly InfluVac Tetra 2022/23 vaccine (Abbott Biologicals B.V., Olst, Netherlands) was administered as most HCWs enrolled to the study were recruited from a single tertiary hospital in Germany. Participation was also open for HCWs from surrounding hospitals and medical surgeries. The detailled study protocol of the CoVacSer study has been described previously. (22)

The Influenza A season 2022/23 in Germany (predominantly H3N2) lasted from 24 October 2022 to 8 January 2023 with its peak occurring in the 50^th^ calendar week of 2022. Subsequently, there was no second Influenza A wave during the 2022/23 winter season. Compared to the pre-pandemic Influenza waves, the 2022/23 seasonal wave occurred considerably earlier, due to the high overall epidemiological ARI activity at the time the COVID-19 pandemic restriction measures ware phased out. Other ARI pathogens, including SARS-CoV-2, RSV, hCoV, hMPV, Parainfluenza virus, and Rhinovirus, were also circulating to varying extents. (3, 23)

The epidemiological estimate of the ARI prevalence in Germany, defined as the clinical syndrome of “acute pharyngitis, bronchitis or pneumonia with or without fever”, is based on virological surveillance infrastructure of patients of all ages in nationwide installed sentinel surgeries. The period of the Influenza A season (Influenza wave) is determined based on the Influenza A positive rate of patient samples obtained through these surveillance structures as follows: (3, 23)

1. Beginning: The lower limit of the 95% confidence interval of the estimated Influenza positive rate in two consecutive calendar weeks exceeds 10%.
2. Ending: The lower limit of the confidence interval of the positive rate falls below 10% in two consecutive weeks with the week before falling below 10%.

### 2.2 Data collection

The data was collected between the 23 May 2022 to the 11 May 2023. Per participation, the study participants submitted the CoVacSer study survey with a corresponding serum blood sample. The date of serum sample receipt was defined as the date of participation.

As a preliminary investigation, we first evaluated whether the administration of the seasonal tetravalent Influenza vaccine affects the obtained levels of Anti-Influenza-A-Nucleoprotein/Matrix IgG in a subgroup of the total study population. This effect was not expected due to the specific target antigens not present in common intramuscular Influenza vaccines (Influenza A Nucleoprotein and Matrix), (2) and the evaluation was conducted based on an independent assessment of the ELISA used in the study. The following two time points were chosen and compared:

- **“pre-vaccination” participation:** Latest study participation ≤ 100 days before the administration of the 2022/23 seasonal tetravalent Influenza vaccination
- **“post-vaccination” participation:** First study participation from 14 up to 40 days after the 2022/23 seasonal tetravalent Influenza vaccination

Based on this preliminary assessment, data was collected and comparatively analysed at the following two time points to determine Influenza A seroconversion and its determinants as the main analysis.

1. **Pre-seasonal data collection**: Latest study participation between 23 May 2022 (defined based on the end of the last Influenza A wave in the 2021/22 Influenza season in Germany), and 23 October 2023. (3)
2. **Post-seasonal data collection**: First study participation between 9 January 2023, and 11 May 2023 (end of the data collection period).

Only serum samples with an attached signed consent form and a completed linked questionnaire were considered. The CoVacSer study questionnaire is composed of socio-demographic characteristics and individual risk factors. The technological platform used for questionnaire collection was REDCap (Research Electronic Data Capture, projectredcap.org). (24) Conditional on pseudonymisation, blood samples were matched to the study survey based on date of birth and COVID-19 vaccination data.

### 2.3 Anti-Influenza-A IgG ELISA and seroconversion

Anti-Influenza-A-Nucleoprotein/Matrix IgG levels were determined using the SERION ELISA classic Influenza A IgG assay (SERION diagnostics, Würzburg, Germany), an enzyme-linked immunoassay. The obtained extinction values were converted to manufacturer-specific Serion IgG Units per ml (U/ml) using the easyANALYZE software (SERION diagnostics).

Infection rate was defined as increase of Anti-Influenza-A-Nucleoprotein/Matrix IgG by a factor of ≥2 since a previous study has modelled that this most closely corresponds to the actual infection rate. (13)

### 2.4 Ethical approval

The study protocol was approved by the Ethics committee of the University of Würzburg in accordance with the Declaration of Helsinki (file no. 79/21).

### 2.5 Statistics

Data analysis was performed using GraphPad Prism 10.0.0 (GraphPad Software, San Diego, CA, USA).

Pairwise comparisons of Anti-Influenza-A-Nucleoprotein/Matrix IgG levels before and after the seasonal Influenza vaccination, before and after the 2022/23 Influenza A season, as well as the factor of Anti-Influenza-A-Nucleoprotein/Matrix IgG level seroconversion, were conducted using the Wilcoxon rank test.

For the univariate comparative distribution analysis of the two cohorts (no seasonal vs. seasonal Influenza vaccination as well as seroconversion vs. no seroconversion; *Table 2*), Fisher’s exact test was used for the binary variables (gender distribution, smoking, regular patient contact, seasonal Influenza vaccination and SARS-CoV-2 infection during the Influenza A season) and Chi square test for the comparison of occupational groups. For the ordinal variables (age, BMI, household members) the Mann Whitney U test was used. To correct against multiple testing, the resulting p-values were adjusted using the Benjamini-Yekutieli procedure (25). The two-tailed significance level α was set to 0.05. Confidence intervals were calculated using the Wilcoxon-Brown Method. (26)

A multiple logistic regression model was employed to evaluate the effect of gender, age, BMI, smoking, members living in the same household, regular patient contact, seasonal Influenza vaccination, and SARS-CoV-2 infection during the Influenza A season on Influenza A seroconversion.

The ordinal variables age, BMI, and household size were dichotomised for the regression analysis. Age was categorized into two groups: individuals under 45 years old and those aged 45 years or older. Household size was divided into two groups: households with fewer than 3 members and households with 3 members or more. The thresholds were chosen to divide the cohort into groups of approximately equal size (*Table 2*). Regarding BMI, two groups were established: individuals with a BMI under 30 kg/m^2^ and individuals with a BMI of 30 kg/m^2^ or higher, following the definition of obesity.

Vaccine effectiveness on seroconversion (VE) of the 2022/23 seasonal Influenza vaccine was calculated using the odds ratio obtained from the multiple logistic regression. The serological VE was derived based on the incidence rate ratio (IRR), defined as the ratio of the probability (p) of seroconversion in the vaccinated subgroup to that in the unvaccinated subgroup using the following formula: (27)

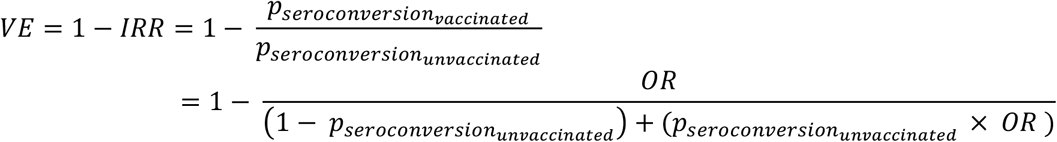

The 95% confidence intervals of the serological VE were calculated using the same method, utilizing the corresponding confidence intervals of the odds ratio derived from the multiple logistic regression analysis. (27)

## 3 Results

### 3.1 Participant recruitment and characterisation of the study population

From 23 May 2022 to 11 May 2023, 548 CoVacSer study participants were enrolled with 1,280 participations and Anti-Influenza-A-Nucleoprotein/Matrix IgG levels were determined.

For the analysis of the test inference of the Anti-Influenza-A-Nucleoprotein/Matrix IgG ELISA and the seasonal tetravalent Influenza vaccine, 37 study participants with paired pre- and post-vaccination participation were included according to the study protocol. 479 individuals could not be included because they had not received the seasonal Influenza vaccine, did not provide information on the date of vaccination, or participated out of the 14 to 40 days post-vaccination interval. A further 32 individuals did not participate prior vaccination or were vaccinated too long ago (*Figure 1*).

**Figure 1:**
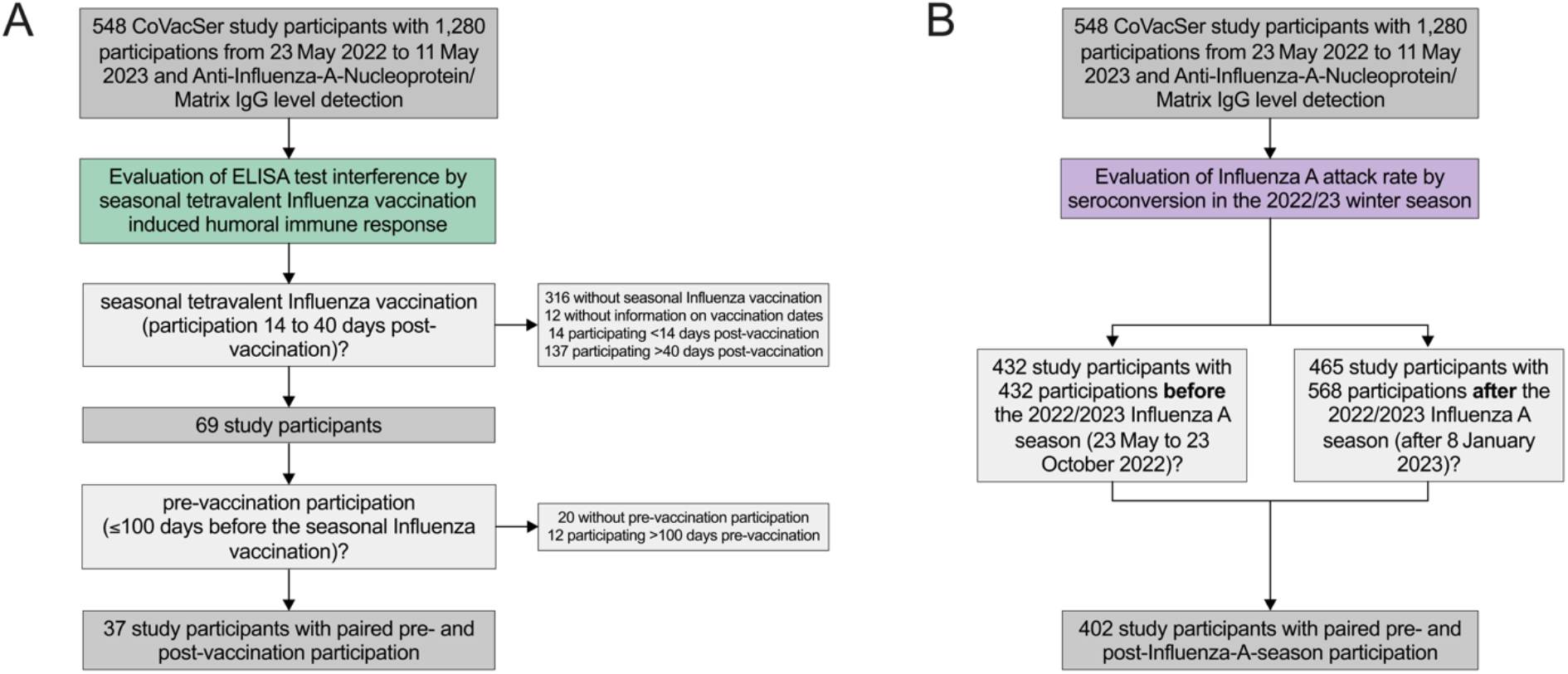
Recruitment of study participants. A) Evaluation of test inference by the seasonal Influenza vaccination B) Evaluation of the Influenza A infection rate and its influencing factors

In the main analysis, the Influenza A seroconversion assessment, 432 individuals participated exactly once from 23 May to 23 October 2022 as a pre-season assessment. After the Influenza season after 8 January 2023, 465 individuals with 568 participations were available. In total this resulted in 402 enrolled individuals with paired pre- and post-season participation (*Figure 1*).

### 3.2 Influence of seasonal Influenza vaccination on Anti-Influenza-A-Nucleoprotein/Matrix IgG

To evaluate interference between the used Anti-Influenza-A-Nucleoprotein/Matrix IgG ELISA and the seasonal Influenza vaccination, IgG titres before and after seasonal Influenza vaccination were compared in a sub-cohort (*Figure 2*). This cohort is characterised in *Table 1*.

**Table 1:**
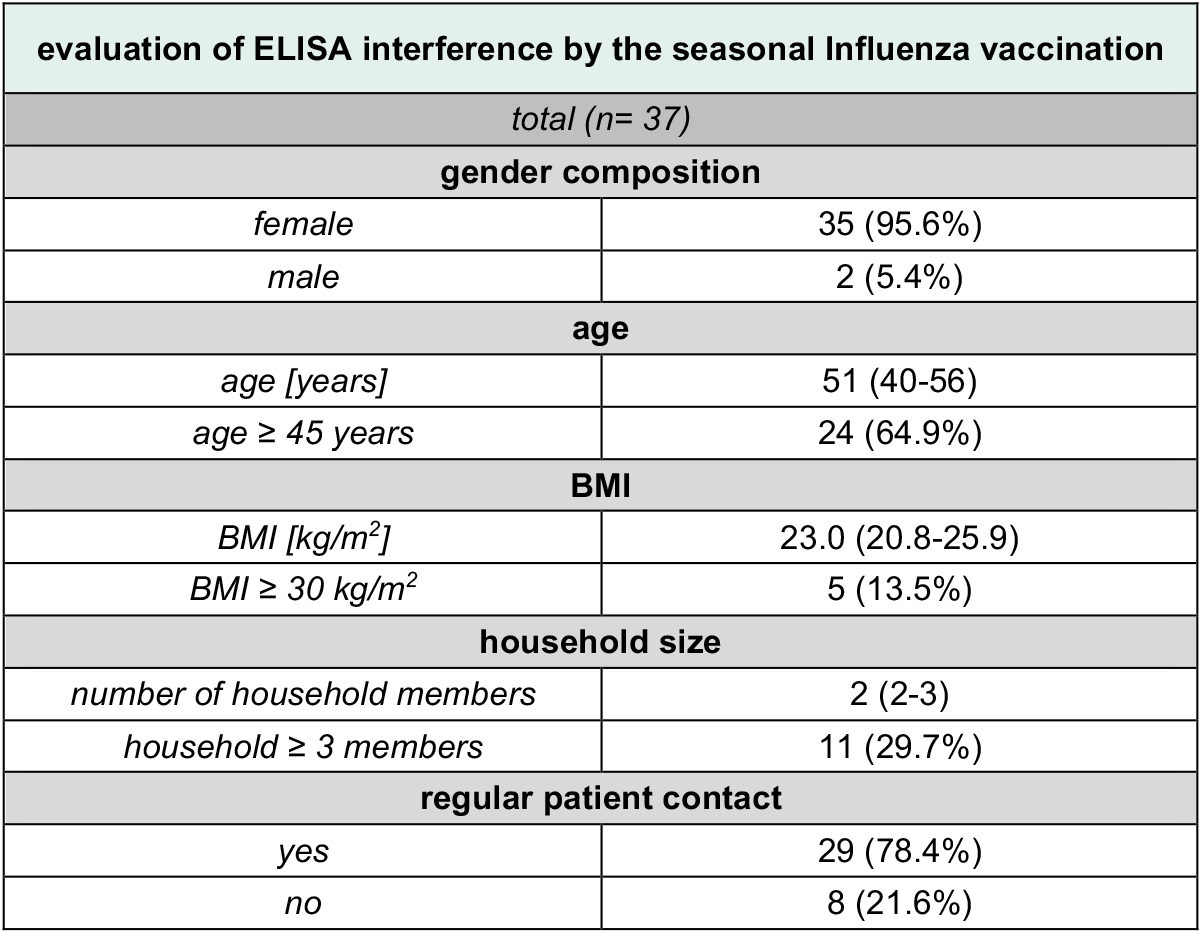
Characterisation of the study population for the preliminary analysis Age [years], BMI [kg/m^2^], and number of household members are given as medians with interquartile ranges in brackets. All other characteristics are presented as absolute numbers with the respective relative numbers in relation to the total cohort in brackets.

**Figure 2:**
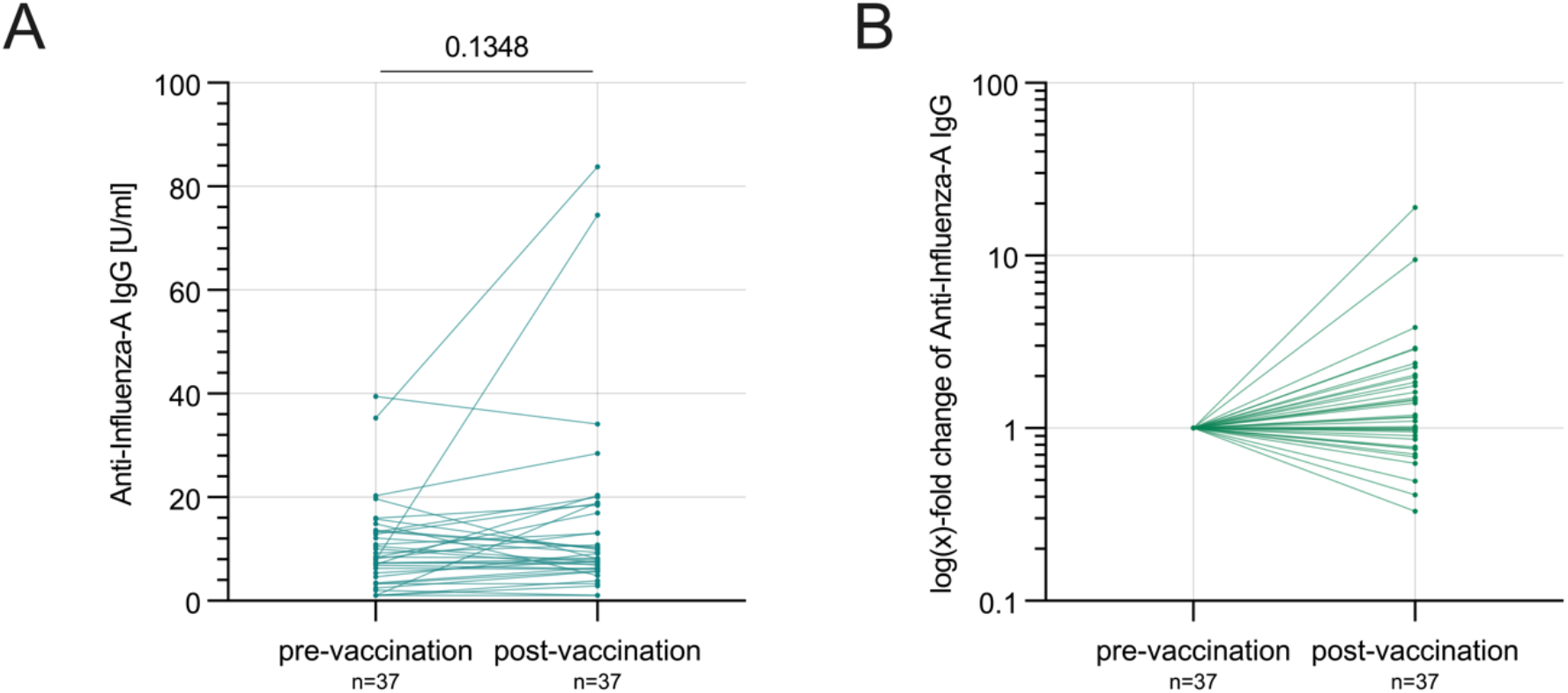
Anti-Influenza-A-Nucleoprotein/Matrix IgG before and after the seasonal Influenza vaccination. A) Anti-Influenza-A-Nucleoprotein/Matrix IgG levels in U/ml before and after the seasonal Influenza vaccination (absolute numbers) B) Relative change of Anti-Influenza-A-Nucleoprotein/Matrix IgG levels after the seasonal Influenza vaccination compared to the pre-vaccination assessment as baseline reference

Median Anti-Influenza-A-Nucleoprotein/Matrix IgG before vaccination was 8.1 (IQR: 4.0-13.4) U/ml, and in the interval 14 to 40 days after vaccination 8.2 (IQR: 6.2-17.7) U/ml. No significant difference was observed in the pairwise comparison (p=0.13; *Figure 2*).

### 3.3 Anti-Influenza-A-Nucleoprotein/Matrix IgG and seroconversion before and after the 2022/23 winter season

Anti-Influenza-A-Nucleoprotein/Matrix IgG levels were assessed before and after the Influenza A 2022/23 season in Germany in a cohort of 402 enrolled individuals. This cohort is characterised in general and stratified by seroconversion in *Table 2*.

**Table 2:**
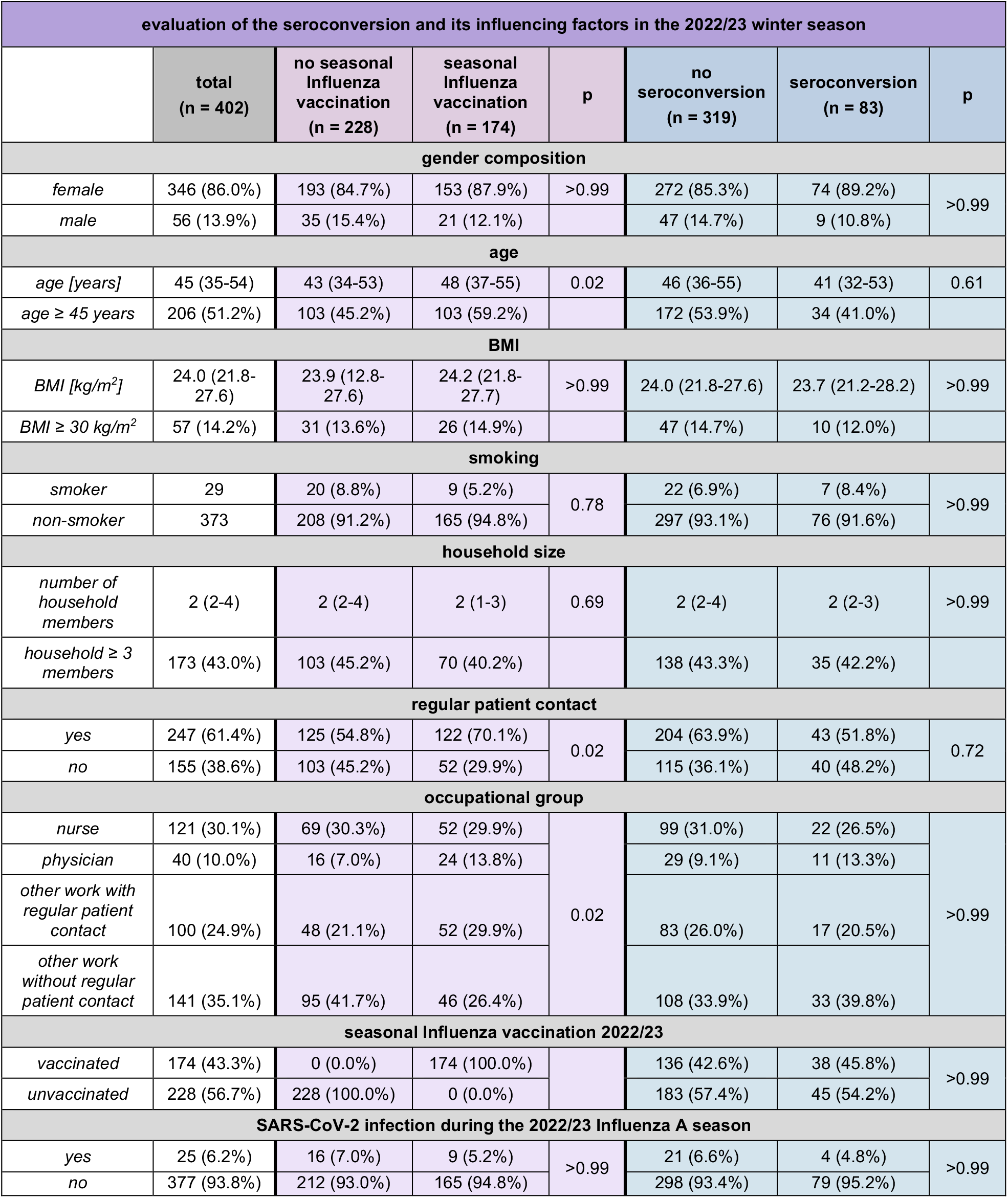
Characterisation of the study population separated by seasonal Influenza vaccination and seroconversion Age [years], BMI [kg/m^2^], and number of household members are given as medians with interquartile ranges in brackets. All other characteristics are presented as absolute numbers with the respective relative numbers in relation to the total cohort in brackets.

Anti-Influenza-A-Nucleoprotein/Matrix IgG levels were 6.0 (IQR: 3.3-10.3) U/ml at median before the 2022/23 Influenza A season and increased by 18.6% (median 7.1; IQR: 4.3-13.2 U/ml) on average after the 2022/23 Influenza A winter season (*Figure 3*).

**Figure 3:**
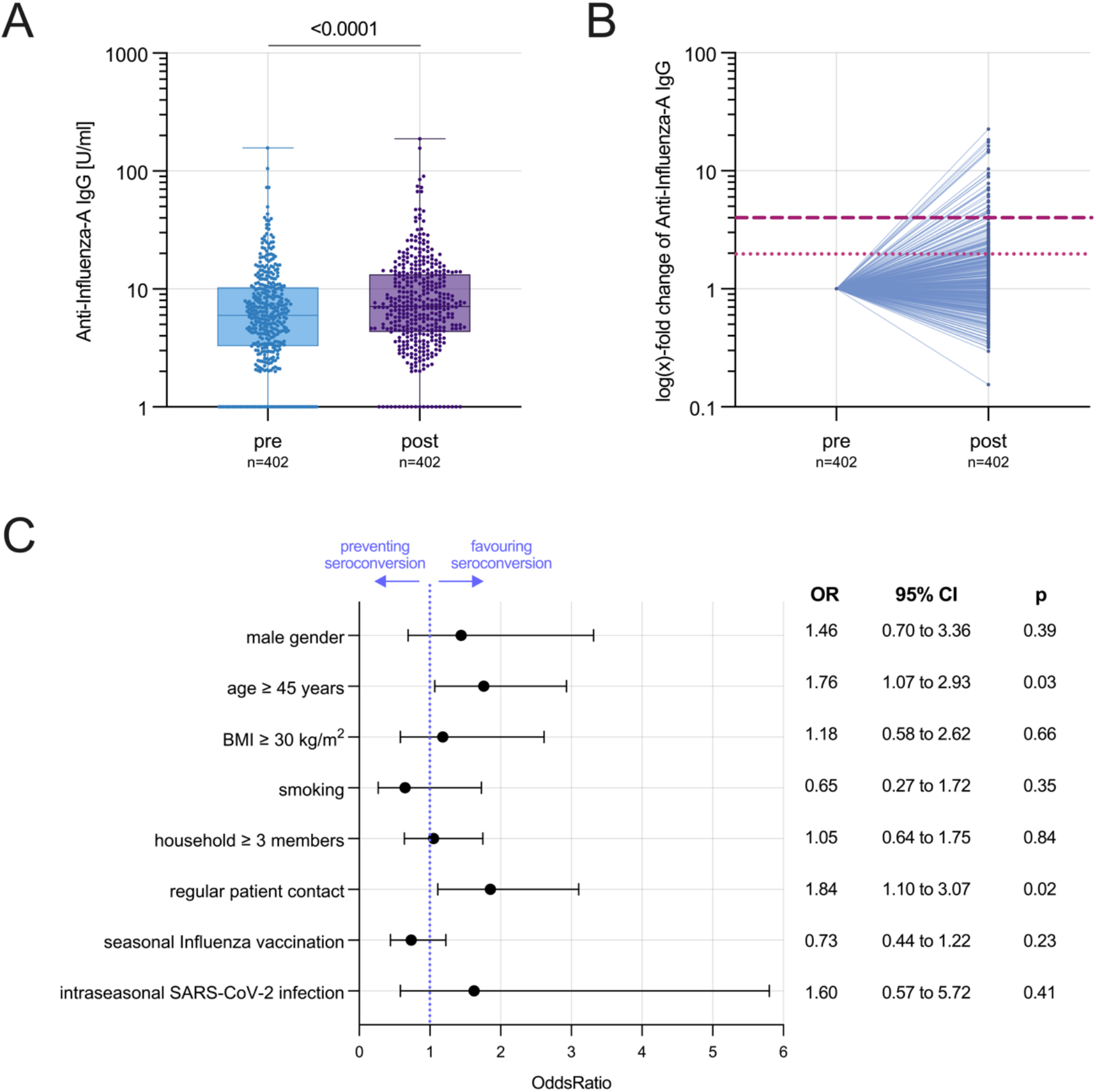
Anti-Influenza-A-Nucleoprotein/Matrix IgG seroconversion and its influencing factors. A) Anti-Influenza-A-Nucleoprotein/Matrix IgG levels in U/ml before and after the 2022/23 Influenza A season B) relative change of Anti-Influenza-A-Nucleoprotein/Matrix IgG levels after the 2022/23 Influenza A season compared to the pre-season assessment as baseline reference C) Odds Ratio of potential factors favouring or preventing Influenza A seroconversion during the 2022/23 Influenza A winter season. Seroconversion was defined as at least twofold increase of the post-season Anti-Influenza-A-Nucleoprotein/Matrix IgG level compared to the pre-season assessment. Error bars indicate 95% CI.

In 20.6% (95% CI 17.0-24.9%; 83/402) of HCWs, there was an antibody level increase by a factor ≥ 2 and thus a seroconversion according to the definition. In 37.3% (150/402, there was an increase in antibody levels by a factor < 2. No increase in Anti-Influenza-A-Nucleoprotein/Matrix IgG was obtained in 42.0% (169/402), antibody levels thus decreased or remained the same during the Influenza season.

In the group of seroconverted HCWs, 60.2% (50/83) showed an increase in IgG titres in the factor range between 2 and 4-fold, 19.3% (16/83) between 4 and 6-fold, 10.8% (9/83) between 6 and 10-fold, and 9.6% (8/83) ≥ 10-fold. The maximum increase in IgG levels was observed by a factor of 22.6 (*Figure 3*).

### 3.4 Seasonal Influenza vaccination: vaccination acceptance and factors influencing it among HCWs

When comparing the two subcohorts of HCWs - one comprising HCWs who received a seasonal Influenza vaccination and the other consisting of HCWs without seasonal influenza vaccination - significant findings emerged. It was observed that older HCWs (p = 0.02) as well as HCWs with regular patient contact (p = 0.02) were significantly more likely to have received a seasonal influenza vaccination. However, no notable differences were found in terms of BMI, household size, smoking, or SARS-CoV-2 infection during the Influenza A season (*Table 2*).

### 3.5 Factors influencing Anti-Influenza-A-Nucleoprotein/Matrix IgG seroconversion

The univariate analysis comparing the two groups obtained no significant differences between the non-seroconverted and the seroconverted HCWs (*Table 2*).

In the multiple regression analysis, regular patient contact (p = 0.02) and age ≥ 45 years (p = 0.03) showed a significant positive correlation with seroconversion. Further, male gender, obesity, ≥ 3 household members, and a SARS-CoV-2 infection during the 2022/23 Influenza A season emerged as factors favouring seroconversion without statistical significance in the regression model. Smoking and seasonal Influenza vaccination were identified as factors that counteracted seroconversion but were also not statistically significant. The Odds Ratio (OR) including the corresponding 95% CIs as well as the p-values are visualised in *Figure 3*.

The VE of the seasonal tetravalent Influenza vaccination determined from the multiple regression analysis was 22.6% (95% CI -17.1 – 50.6%).

## 4 Discussion

In the first Influenza A season following the COVID-19 pandemic, about one fifth of HCWs contracted Influenza A, which highlights a concerning occupational infection rate. The Anti-Influenza-A-Nucleoprotein/Matrix IgG titres increased significantly after the end of the Influenza A wave compared to the pre-pandemic assessment as baseline reference, so that the epidemiological, symptom-based infection detection could also be reproduced serologically. (3)

The infection rate significantly correlated with older age as well as regular patient exposure. The latter is especially interesting as face masking was still common in the German healthcare sector during large parts of the study period. Intraseasonal SARS-CoV-2 infection, which can be seen as an indicator for exposure to respiratory pathogens as well as risk behaviour that may lead to an Influenza A infection, e.g. not wearing a face mask or wearing it ineffectively, was positively correlated with Influenza A infection rate although the effect was not statistically significant. In contrast to all other ARI pathogens including Influenza A, diagnostics for SARS-CoV-2 were available at any time during the entire 2022/23 winter season by means of PCR or rapid antigen tests for respiratory symptoms, which further emphasises the genuineness of this predictive factor.

The effectiveness of the seasonal, tetravalent Influenza vaccination, taking into account seroconversion and thus the potential underreporting, was 26.3%, which falls below the range of vaccine effectiveness reported by Kissling et al. in an European interim analysis of the 2022/23 seasonal Influenza vaccination. (28) This can be explained by the methodological difference: While the data presented is based on seroconversion the analysis of Kissling et al. was based on symptoms and PCR confirmed cases of Influenza A. Previous evidence demonstrated clearly that the purpose of the seasonal Influenza vaccination is not only to prevent infection, but also to mitigate the clinical course so that the infection is only mildly symptomatic or completely asymptomatic in the event of infection. (29) Consequently, in the context of an infection or symptom-based vaccine effectiveness analysis, it is precisely those mild or symptomatic infections among vaccinated persons that are not recorded, which, in contrast, is possible in a serologically based evaluation, as carried out here. In this way, undetected infections can also be recorded, which consequently lowers the vaccine effectiveness. (30)

We deliberately used Anti-Influenza-A-Nucleoprotein/Matrix IgG to determine the infection rate instead of the commonly used haemagglutinin inhibition assay. This approach allowed us to accurately detect seroconversion resulting from natural Influenza A infection. While there is no published evidence on test validation for the specific ELISA we used, the data we present now enables such validation for the first time.

In the specificity analysis, conducted on a sub-cohort of the study population, we observed no significant change in Anti-Influenza-A-Nucleoprotein/Matrix IgG levels following seasonal Influenza vaccination. However, a small proportion of individuals experienced an increase in IgG titre after vaccination. It’s important to note that this finding should be interpreted in the context of random Influenza A infections occurring between the two measurement points. Additionally, this proportion aligns with the general infection rate observed in our study.

Other minor fluctuations, including discrete increases in IgG levels from pre-to post-measurement, can be attributed to common serological measurement fluctuations. The evidence on Influenza A serology remains limited, underscoring the need for test validations based on our pioneering work.

The study presented has several limitations that need to be considered when interpreting the data. Over 80% of the study participants are female. This can be explained by the characteristic composition of HCWs in the German healthcare system, with a clear preponderance of female employees. (31) The study analyses only Influenza A infection rate, as Influenza A has a considerably higher disease burden and a pandemic potential compared to the likewise established Influenza B, which also circulates as an ARI pathogen seasonally in Germany. (3, 7) The infection rate is based on serological evaluation which never has perfect sensitivity and specificity. The threshold of a ≥ 2-fold increase was based only on data from one study as no other evidence-based thresholds were available. (13) It was not recorded how often and when the participants had a concrete exposure to Influenza A positive persons during the study period, nor how often and when typical symptoms of Influenza A (Influenza like illness, ILI) occurred. Thus, the study objectively detects the infection rate, but not the symptom-based defined real attack rate, for which further investigations with detailed ILI history assessment during the course of the entire winter season and Influenza A diagnostics in case of any symptoms are necessary. Similarly, it was not recorded whether a seasonal Influenza vaccination was administered in the winters prior to the study period.

The infection rate based on longitudinal antibody testing is about 60-times higher and presumably represents infection rates, including asymptomatic infections, more completely than the reported data of the national acute respiratory infection surveillance for the 2022/23 winter season, which is associated with substantial underdiagnosis and indicates an Influenza infection rate of 0.3% for Germany in the 2022/23 season. (32) However it remains questionable and should be studied in how far persons with asymptomatic Influenza infection resulting in seroconversion contribute to spreading of the virus in the community. Knowledge of the actual infection rate is of great relevance for evaluation and planning of prevention strategies against Influenza, especially vaccination strategies, in both healthcare settings and the general population.

## Data Availability

Additional data that underlie the results reported in this article, after de-identification (text, tables, figures, and appendices) as well as the study protocol, statistical analysis plan, and analytic code is made available to researchers who provide a methodologically sound proposal to achieve aims in the approved proposal on request to the corresponding author.

## Author contributions

All authors had unlimited access to all data. Isabell Wagenhäuser and Manuel Krone take responsibility for the integrity of the data and the accuracy of the data analysis.

*Conception and design*: Isabell Wagenhäuser, Juliane Mees, Julia Reusch, Thiên-Trí Lâm, Alexandra Schubert-Unkmeir, Lukas B. Krone, Anna Frey, Oliver Kurzai, Stefan Frantz, Lars Dölken, Johannes Liese, Alexander Gabel, Nils Petri, Manuel Krone

*Trial management*: Isabell Wagenhäuser, Juliane Mees, Julia Reusch, Alexander Gabel, Nils Petri, Manuel Krone.

*Laboratory analysis:* Isabell Wagenhäuser, Juliane Mees, Julia Reusch.

*Statistical analysis*: Isabell Wagenhäuser, Manuel Krone.

*Obtained funding*: Oliver Kurzai, Manuel Krone.

*First draft of the manuscript*: Isabell Wagenhäuser, Manuel Krone. The manuscript was reviewed and approved by all the authors.

## Acknowledgements

We thank the staff of the serological diagnostic laboratory for making their laboratory available and especially for their advisory help.

We explicitly thank Professor Ulrich Vogel, Infection Control and Antimicrobial Stewardship Unit, University Hospital Würzburg, Germany, for conception and design as well as funding support. He played a major role regarding the CoVacSer study but passed away much too early during the study. We miss him as college and friend who showed a great dedication to his work, family, and friends.

## Statement on the use of artificial intelligence (AI) in the writing process

Artificial intelligence (AI) was used for language improvement purposes only. The tools ChatGPT (OpenAI, San Francisco CA, USA) and DeepL (DeepL SE, Cologne, Germany) were used. The actual writing of the manuscript was carried out solely by the authors mentioned by name.

